# Use of over-the-counter-like drugs among adults with indicated chronic diseases: A nationwide descriptive study in Japan

**DOI:** 10.64898/2026.02.03.26345444

**Authors:** Yuya Kimura, Shotaro Aso, Akira Okada, Hideo Yasunaga

**Author notes:** Correspondence: Yuya Kimura Department of Health Services Research, Graduate School of Medicine, The University of Tokyo, Tokyo, Japan.

## Abstract

**Introduction:** The Japanese Ministry of Health, Labour, and Welfare proposed increased copayments for over-the-counter (OTC)-like drugs, with possible special consideration for certain chronic diseases, although the specific diseases to be included were not clarified as of January 2026. The costs attributable to OTC-like drugs indicated for chronic diseases in adults remain unclear.

**Methods:** Using the DeSC database (individual-level claims data) in fiscal year 2023, we estimated the average per-person annual costs of the study OTC-like drugs indicated for adults with representative chronic diseases. The estimates were stratified by sex and age. We also computed the national estimates of the numbers of adults with representative chronic diseases who would need OTC-like drugs and the costs attributable to these drugs by applying stratum-specific estimates from the DeSC database to national population counts and aggregated claims data from the National Database of Health Insurance Claims Open Data. The study OTC-like drugs included oral acetaminophen, oral nonsteroidal anti-inflammatory drugs (NSAIDs), topical anti-inflammatory patches, oral second-generation antihistamines, and heparinoid-containing topical preparations.

**Results:** The average per-person annual costs were several hundred yen for acetaminophen and several thousand yen for the other drug categories, with marked variation by drug category, chronic disease, age, and sex. In the national estimates, the proportion of OTC-like drug users with representative chronic diseases was <10% in every disease–age–sex stratum; nevertheless, the maximum stratum-specific shares of total OTC-like drug costs attributable to these groups were high (up to 50–60% for osteoarthritis for acetaminophen/NSAIDs/patches and up to ∼60% for atopic dermatitis/asteatosis for heparinoids).

**Conclusions:** The average per-person annual costs for OTC-like drugs varied substantially across drug categories and patient subgroups. Indicated chronic diseases in adults may account for substantial OTC-like drug costs within some strata, despite representing a small fraction of users.

## Introduction

“*Over-the-counter (OTC)-like drugs*” are prescription medications that contain the same or similar ingredients as OTC drugs, which are considered sufficiently safe to be sold without a prescription^1^. In Japan, out-of-pocket costs for OTC-like drugs are generally lower than those for their OTC counterparts because the former are reimbursed under the public health insurance system, requiring patients to pay only a portion of the total costs (typically 10–30%), whereas OTC drugs are paid for fully out of pocket. Furthermore, the prices of OTC drugs often include marketing and distribution costs. Against this background, an active policy debate has been ongoing from 2025 to 2026 in Japan regarding whether OTC-like drugs should be excluded from public health insurance coverage or subject to increased patient copayments, with the aim of reducing national health expenditures and alleviating the financial burden of insurance premiums^2^. In our previous research focusing on representative OTC-like drugs, viz. acetaminophen, nonsteroidal anti-inflammatory drugs (NSAIDs), topical anti-inflammatory patches, second-generation antihistamines, and heparinoid-containing topical preparations, the total prescription volume and costs in fiscal year (FY) 2023 reached 17.4 billion prescriptions (139.6 per capita) and 288.1 billion yen (2,317.0 yen per capita), respectively^1^. We further demonstrated substantial heterogeneity in the use of these drugs according to patient characteristics, including sex, age, and comorbidity burden.

As of December 2025, the Ministry of Health, Labour, and Welfare of Japan (MHLW) reported that OTC-like drugs would not be excluded from public health insurance coverage but would be subject to higher patient copayments^2^. The MHLW indicated that special consideration would be given to certain populations, including children, patients with chronic diseases (e.g., malignant neoplasms and intractable diseases), and patients in whom long-term use of OTC-like drugs was judged to be medically necessary by physicians. However, as of January 2026, the types of chronic diseases, beyond malignant neoplasms and intractable diseases, which would be included in these considerations are unclear. Although adult patients with representative chronic diseases for which OTC-like drugs are indicated may bear a substantial cost burden, evidence regarding the magnitude of the expenses attributable to these drugs in this population is lacking. Therefore, using nationwide claims data and focusing on the same representative OTC-like drugs examined in our previous research^1^, we aimed to quantify the costs attributable to these drugs, which are indicated for adult patients with representative chronic diseases.

## Methods

### Data source

In this study, we used two data sources: 1) the National Database of Health Insurance Claims (NDB) Open Data and 2) the DeSC database. Detailed descriptions of these databases have been provided in our previous research^1^. Briefly, the NDB is a nationwide claims database covering over 99% of healthcare facilities in Japan and includes information on patient demographics (sex and age); diagnoses coded using the International Classification of Diseases, 10^th^ Revision; procedures; and medications, all of which were extracted from both inpatient and outpatient settings^3,4^. The NDB Open Data are publicly available aggregated datasets derived from the NDB and include annual prescription volumes and costs overall and stratified by sex and age group.

The DeSC database is a commercially available claims database whose data elements are similar to those of the NDB^4,5^. While the NDB provides comprehensive coverage of all five public health insurance types in Japan, the DeSC database collects data from three insurance types. Detailed comparisons across the number and proportion of enrollees in each insurance type across both databases have been described elsewhere^1^. We analyzed data in FY 2023 (April 2023 to March 2024) from both data sources.

### Study drugs

We examined the same five categories of representative OTC-like drugs as in our previous research^1^: oral single-agent acetaminophen, oral single-agent NSAIDs, topical anti-inflammatory patches, oral second-generation antihistamines, and heparinoid-containing topical preparations. The analysis was limited to outpatient prescriptions because inpatient drug costs are often bundled into inclusive reimbursement systems, making it difficult to identify costs attributable to individual drugs. The World Health Organization–Anatomical Therapeutic Chemical codes for each drug category are provided in our previous research^1^.

### Costs attributable to OTC-like drugs among adult patients with representative chronic diseases

From the DeSC database, we identified adult patients with representative chronic diseases for which the study OTC-like drugs are indicated, who received prescriptions for these study drugs in FY2023. Representative chronic diseases were considered to be present if the corresponding diagnostic codes were recorded at least once in FY2023. Patient identification was performed within strata defined by the combination of sex (male or female) and age category (18–34, 35–44, 45–54, 55–64, 65–74, 75–84, 85–94, and ≥95 years). Stratification by sex and age group was conducted because the costs attributable to the study drugs in these patient groups were expected to vary substantially across the strata. Age categories were determined to align with major thresholds in the Japanese health insurance system, particularly at ages 65 and 75 years. Based on the number of such patients and the costs assignable to the study drugs, we calculated the average annual per-person costs attributable to the study drugs.

The following representative chronic diseases for which each study drug is generally indicated were identified: acetaminophen, NSAIDs, and topical anti-inflammatory patches for osteoarthritis and rheumatoid arthritis; second-generation antihistamines for allergic rhinitis, urticaria, and atopic dermatitis; and heparinoid-containing topical preparations for the combination of atopic dermatitis and asteatosis. The International Classification of Diseases, 10^th^ Revision codes for each disease are as follows: M16–17 for osteoarthritis, M05–06 for rheumatoid arthritis, J30 for allergic rhinitis, L50 for urticaria, L20 for atopic dermatitis, and L853 for asteatosis.

### Estimates of the national number of adult patients with representative chronic diseases who would need OTC-like drugs and the costs attributable to these drugs

The supplementary analyses entailed estimation of the number of adult patients nationwide with representative chronic diseases who would need the study OTC-like drugs and the corresponding national costs attributable to these drugs. This estimation was conducted using a two-step approach. First, using data from the DeSC database, we calculated two stratum-specific proportions within the above-mentioned strata defined by the sex and age category in FY2023: 1) the proportion of adult patients with representative chronic diseases who used the study drugs among all individuals within each stratum; and 2) the proportion of prescription costs attributable to the study drugs in these patients among the total prescription costs within each stratum. Second, the stratum-specific estimates of the first proportion (number-based) were applied to the corresponding sex-and age-specific national population counts obtained from the Statistics Bureau of Japan to estimate the national number of adult patients with representative chronic diseases who would need the study OTC-like drugs^6^. Furthermore, the stratum-specific estimates of the second proportion (cost-based) were applied to the corresponding stratum-specific prescription costs in the NDB Open Data to derive the national cost estimates of the study drugs.

## Results

### Costs attributable to OTC-like drugs among adult patients with representative chronic diseases

Among adults with representative chronic diseases who received prescriptions for the study OTC-like drugs in FY2023, the average per-person annual costs by sex and age category are presented in Tables 1–5 and Figure 1.

**Figure 1.**
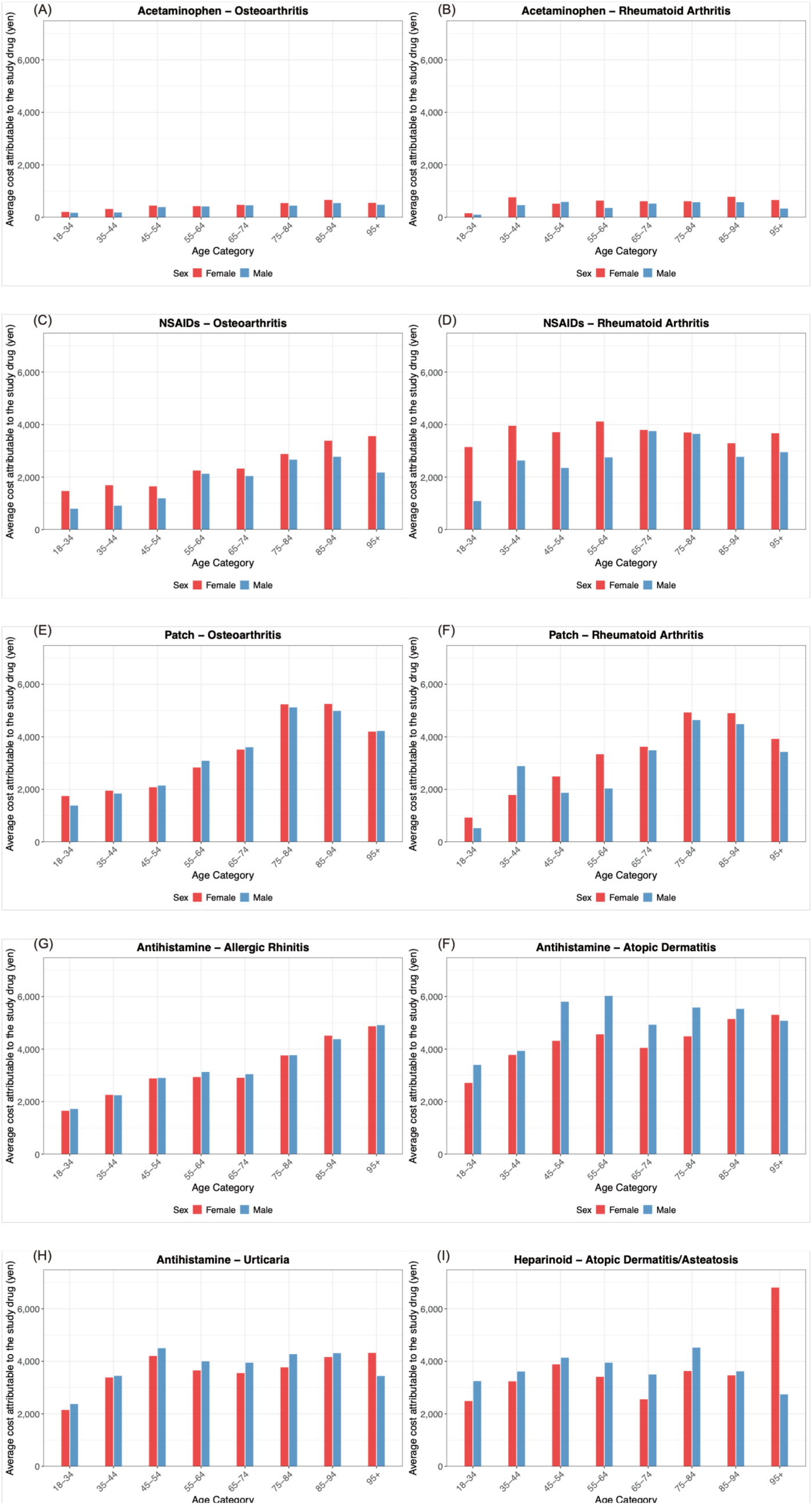
Average per-person annual costs for study OTC-like drugs among adults with representative chronic diseases, by age and sex in the DeSC database. The panels show the average per-person annual costs (Japanese yen) attributable to each study OTC-like drug among adults with the corresponding representative chronic diseases in FY2023, stratified by age and sex. (A) Oral single-agent acetaminophen—osteoarthritis (B) Oral single-agent acetaminophen—rheumatoid arthritis (C) Oral single-agent NSAIDs (nonsteroidal anti-inflammatory drugs)—osteoarthritis (D) Oral single-agent NSAIDs—rheumatoid arthritis (E) Topical anti-inflammatory patches—osteoarthritis (F) Topical anti-inflammatory patches—rheumatoid arthritis (G) Oral second-generation antihistamines—allergic rhinitis (H) Oral second-generation antihistamines—atopic dermatitis (I) Oral second-generation antihistamines—urticaria (J) Heparinoid-containing topical preparations—atopic dermatitis/asteatosis. OTC: over-the-counter, NSAID: non-steroidal anti-inflammatory drug

**Table 1.** Average annual costs for acetaminophen among adults with osteoarthritis and rheumatoid arthritis in the DeSC database.

| Disease and Age Category | Female | Male |
| --- | --- | --- |
| Osteoarthritis |  |  |
| 18–34 | 210.4 | 175.4 |
| 35–44 | 320.1 | 187.5 |
| 45–54 | 449.8 | 393.3 |
| 55–64 | 428.8 | 416.4 |
| 65–74 | 477.4 | 461.3 |
| 75–84 | 545.6 | 448.1 |
| 85–94 | 665.9 | 545.1 |
| ≥95 | 554.0 | 481.5 |
| Rheumatoid arthritis |  |  |
| 18–34 | 158.0 | 102.4 |
| 35–44 | 765.6 | 467.3 |
| 45–54 | 522.3 | 589.3 |
| 55–64 | 639.9 | 360.4 |
| 65–74 | 615.7 | 526.5 |
| 75–84 | 615.2 | 578.0 |
| 85–94 | 785.1 | 578.3 |
| ≥95 | 661.8 | 336.1 |

Costs are expressed in Japanese yen.

**Table 2.** Average annual costs for NSAIDs among adults with osteoarthritis and rheumatoid arthritis in the DeSC database.

| Disease and Age Category | Female | Male |
| --- | --- | --- |
| **Osteoarthritis** |  |  |
| 18–34 | 1471.1 | 797.0 |
| 35–44 | 1692.0 | 912.7 |
| 45–54 | 1648.4 | 1192.7 |
| 55–64 | 2250.7 | 2128.1 |
| 65–74 | 2325.5 | 2041.5 |
| 75–84 | 2881.4 | 2664.4 |
| 85–94 | 3385.9 | 2774.9 |
| ≥95 | 3561.6 | 2175.5 |
| **Rheumatoid arthritis** |  |  |
| 18–34 | 3146.1 | 1089.1 |
| 35–44 | 3954.9 | 2632.7 |
| 45–54 | 3710.5 | 2350.4 |
| 55–64 | 4117.9 | 2751.3 |
| 65–74 | 3797.6 | 3750.8 |
| 75–84 | 3699.7 | 3644.9 |
| 85–94 | 3290.1 | 2772.6 |
| ≥95 | 3669.9 | 2949.7 |

Costs are expressed in Japanese yen.
NSAIDs: nonsteroidal anti-inflammatory drugs

**Table 3.**
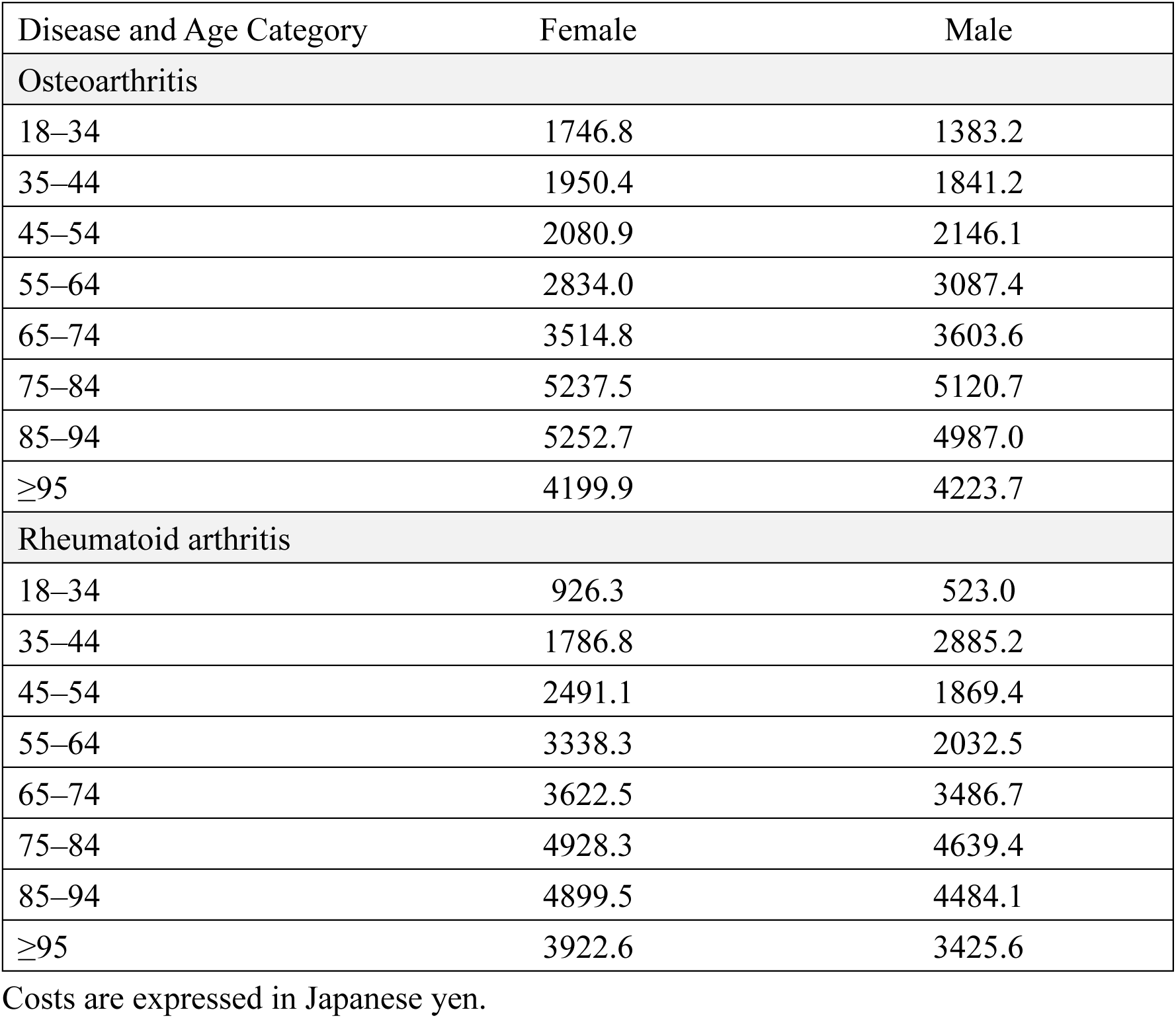
Average annual costs for topical anti-inflammatory patches among adults with osteoarthritis and rheumatoid arthritis in the DeSC database.

**Table 4.** Average annual costs for second-generation antihistamines among adults with allergic rhinitis, urticaria, and atopic dermatitis in the DeSC database.

| Disease and Age Category | Female | Male |
| --- | --- | --- |
| Allergic rhinitis |  |  |
| 18–34 | 1651.8 | 1721.6 |
| 35–44 | 2257.1 | 2242.1 |
| 45–54 | 2881.8 | 2901.7 |
| 55–64 | 2936.1 | 3128.1 |
| 65–74 | 2909.1 | 3044.9 |
| 75–84 | 3759.2 | 3767.3 |
| 85–94 | 4510.9 | 4377.0 |
| ≥95 | 4868.0 | 4911.8 |
| Urticaria |  |  |
| 18–34 | 2148.2 | 2374.3 |
| 35–44 | 3385.0 | 3447.3 |
| 45–54 | 4204.0 | 4500.9 |
| 55–64 | 3654.1 | 4000.3 |
| 65–74 | 3552.7 | 3948.2 |
| 75–84 | 3771.0 | 4272.3 |
| 85–94 | 4163.0 | 4310.4 |
| ≥95 | 4321.4 | 3442.2 |
| Atopic dermatitis |  |  |
| 18–34 | 2711.5 | 3398.9 |
| 35–44 | 3778.5 | 3931.4 |
| 45–54 | 4314.2 | 5803.4 |
| 55–64 | 4560.9 | 6025.5 |
| 65–74 | 4046.2 | 4928.1 |
| 75–84 | 4485.7 | 5581.4 |
| 85–94 | 5145.0 | 5530.7 |
| ≥95 | 5304.0 | 5077.0 |

Costs are expressed in Japanese yen.

**Table 5.**
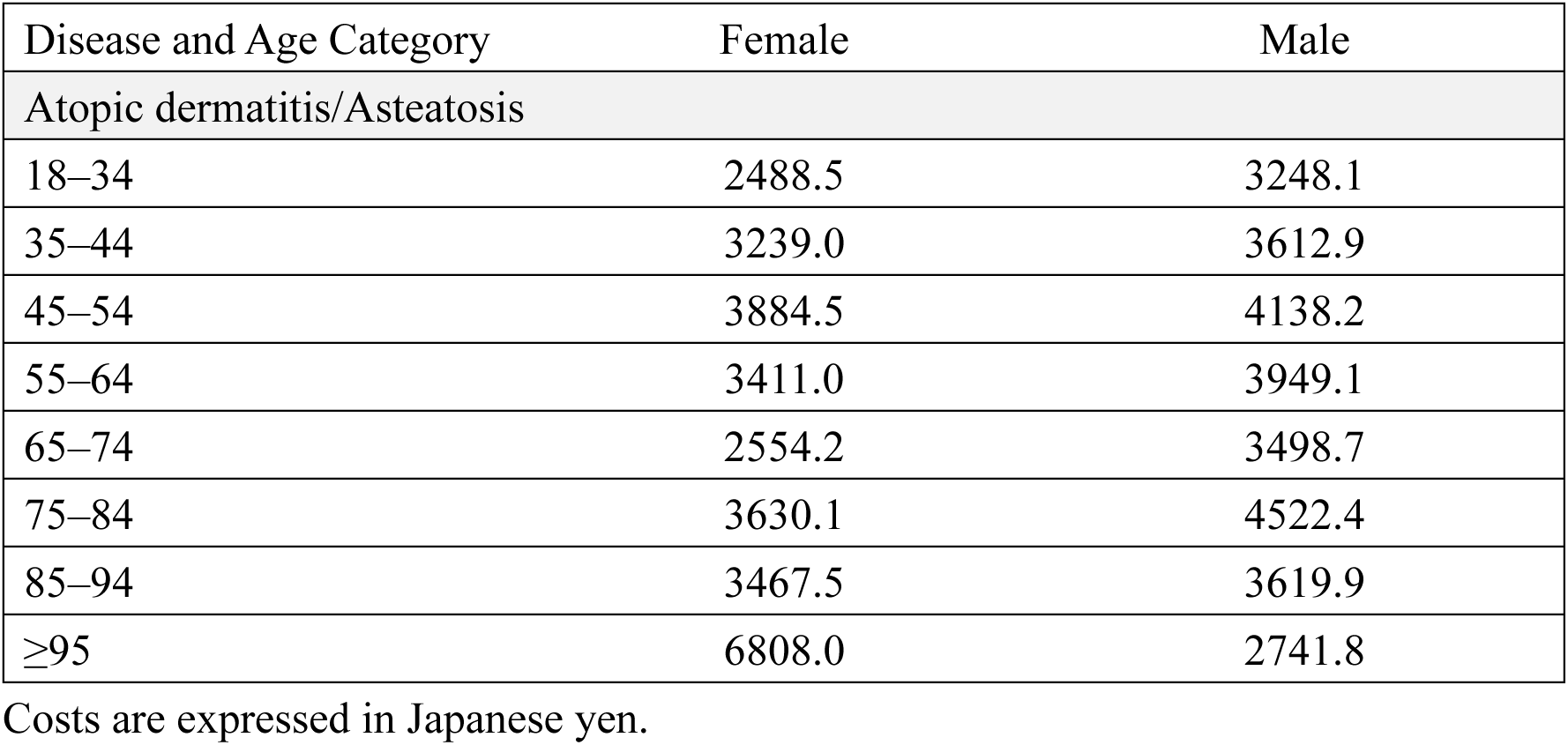
Average annual costs for heparinoid-containing topical preparations among adults with the combination of atopic dermatitis and asteatosis in the DeSC database.

| Disease and Age Category | Female | Male |
| --- | --- | --- |
| Atopic dermatitis/Asteatosis |  |  |
| 18–34 | 2488.5 | 3248.1 |
| 35–44 | 3239.0 | 3612.9 |
| 45–54 | 3884.5 | 4138.2 |
| 55–64 | 3411.0 | 3949.1 |
| 65–74 | 2554.2 | 3498.7 |
| 75–84 | 3630.1 | 4522.4 |
| 85–94 | 3467.5 | 3619.9 |
| ≥95 | 6808.0 | 2741.8 |

Costs are expressed in Japanese yen.

The average per-person annual costs for oral single-agent acetaminophen lay in the several hundred yen range (Table 1 and Figure 1). For osteoarthritis, the costs of this drug among females ranged from 210.4 (aged 18–34 years) to 665.9 yen (aged 85–94 years), while those among males ranged from 175.4 (aged 18–34 years) to 545.1 yen (aged 85–94 years). For rheumatoid arthritis, the costs for single-agent acetaminophen among females ranged from 158.0 (aged 18–34 years) to 785.1 yen (aged 85–94 years), whereas those among males ranged from 102.4 (aged 18–34 years) to 589.3 yen (aged 45–54 years).

For oral single-agent NSAIDs, the average per-person annual costs ranged from approximately 797 to 4118 yen (Table 2 and Figure 1). When prescribed for osteoarthritis, the costs exhibited a categorical age-related increase: among females, the costs rose from 1471.1 yen (aged 18–34 years) to a maximum of 3561.6 yen (aged ≥95 years), whereas among males, they increased from 797.0 yen (aged 18–34 years) to a maximum of 2774.9 yen (aged 85–94 years). When oral single-agent NSAIDs were prescribed for rheumatoid arthritis, no age-related increase was apparent; among females, the costs ranged from 3146.1 (aged 18–34 years) to 4117.9 yen (aged 55–64 years), and from 1089.1 (aged 18–34 years) to 3750.8 yen (aged 65–74 years) among males.

For topical anti-inflammatory patches, the average per-person annual costs ranged from approximately 523 to 5253 yen and increased with age when prescribed for both osteoarthritis and rheumatoid arthritis (Table 3 and Figure 1). For osteoarthritis, the costs ranged from 1746.8 (aged 18–34 years) to 5252.7 yen (aged 85–94 years) across the female strata, and from 1383.2 yen (aged 18–34 years) to 5120.7 yen (aged 75–84 years) across the male strata. For rheumatoid arthritis, costs ranged from 926.3 (aged 18–34 years) to 4928.3 yen (aged 75–84 years) across the female strata, and from 523.0 (aged 18–34 years) to 4639.4 yen (aged 75–84 years) across the male strata.

The average per-person annual costs for oral second-generation antihistamines ranged from approximately 1652 to 6026 yen (Table 4 and Figure 1). For allergic rhinitis, the costs generally increased with age, ranging from 1651.8 (aged 18–34 years) to 4868.0 yen (aged ≥95 years) among females and from 1721.6 (aged 18–34 years) to 4911.8 yen (aged ≥95 years) among males. For urticaria, the costs increased from the 18–34 year group to the 45–54 year group and were generally similar thereafter; among females, the costs ranged from 2148.2 (aged 18–34 years) to 4321.4 yen (aged ≥95 years), and from 2374.3 (aged 18–34 years) to 4500.9 yen (aged 45–54 years) among males. The drug costs for atopic dermatitis increased from the 18–34 year group to the 45–54 year group and were generally similar thereafter; among females, the costs ranged from 2711.5 (aged 18–34 years) to 5304.0 yen (aged ≥95 years), and from 3398.9 (aged 18–34 years) to 6025.5 yen (aged 55–64 years) among males.

For heparinoid-containing topical preparations, the average per-person annual costs ranged from approximately 2489 to 6808 yen (Table 5 and Figure 1), and no clear age-related pattern emerged. Among females, the costs ranged from 2488.5 (aged 1834 years) to 6808.0 yen (aged ≥95 years), and from 2741.8 (aged ≥95 years) to 4522.4 yen (aged 75–84 years) among males.

### National estimates of the number of adult patients with representative chronic diseases who would need OTC-like drugs and the costs attributable to these drugs

We derived national estimates of the proportion and number of adults with representative chronic diseases who would need OTC-like drugs and the costs attributable to these drugs using the age- and sex-specific proportions of adults with representative chronic diseases who used the study OTC-like drugs (Supplementary Tables 1–5) and age- and sex-specific shares of costs attributable to these drugs estimated from the DeSC database (Supplementary Tables 6–10). The resulting national estimates are shown in Supplementary Tables 11–15 (numbers) and 16–20 (costs).

The estimated national number of adults who would need oral single-agent acetaminophen was 0.71 million among those with osteoarthritis and 0.13 million among those with rheumatoid arthritis (Supplementary Table 11). The estimated age–sex stratum proportions fell below 5% for both diseases, the maximum being 2.9% among females aged 85–94 years with osteoarthritis. The estimated national costs for acetaminophen among adults with the indicated chronic diseases were 3.6 billion yen for osteoarthritis and 0.8 billion yen for rheumatoid arthritis (Supplementary Table 16). The maximum cost shares attributable to acetaminophen within specific age–sex strata were 56.0% for osteoarthritis and 10.2% for rheumatoid arthritis.

The estimated number of adults nationwide who would need oral single-agent NSAIDs was 0.54 million among those with osteoarthritis and 0.09 million among those with rheumatoid arthritis (Supplementary Table 12), with age–sex stratum proportions below 5% for both diseases. The estimated national costs attributable to NSAIDs were 6.7 billion yen for osteoarthritis and 1.6 billion yen for rheumatoid arthritis (Supplementary Table 17). The largest cost shares were attributed to osteoarthritis (up to 64.6%) and rheumatoid arthritis (up to 20.9%) within specific age–sex strata.

The estimated number of adults nationwide who would need topical anti-inflammatory patches was 1.84 million among patients with osteoarthritis and 0.22 million among those with rheumatoid arthritis (Supplementary Table 13). The maximum estimated age–sex stratum proportion was 7.0% among females with osteoarthritis aged 75–84 years. The estimated national costs attributable to topical anti-inflammatory patches were 38.2 billion yen for osteoarthritis and 4.4 billion yen for rheumatoid arthritis (Supplementary Table 18). The largest cost shares were ascribed to osteoarthritis (up to 60.8%) and rheumatoid arthritis (up to 8.2%) within specific age–sex strata.

The estimated national number of adults who would need oral second-generation antihistamines was 2.87 million among those with allergic rhinitis, 0.53 million among those with urticaria, and 0.43 million among those with atopic dermatitis (Supplementary Table 14). The maximum estimated age–sex stratum proportion was 3.8% among females with allergic rhinitis aged 75–84 years. The estimated national costs attributable to oral second-generation antihistamines were 71.6 billion yen for allergic rhinitis, 17.3 billion yen for urticaria, and 17.8 billion yen for atopic dermatitis (Supplementary Table 19). The largest cost shares were apportioned to allergic rhinitis (up to 78.5%), urticaria (up to 23.3%), and atopic dermatitis (up to 37.3%) within specific age–sex strata.

The estimated national number of adults who would need heparinoid-containing topical preparations was 0.31 million (Supplementary Table 15). The estimated age–sex stratum proportions were below 1%. The estimated national costs attributable to heparinoid-containing topical preparations were 11.2 billion yen (Supplementary Table 20). The largest cost shares (up to 58.2%) were observed in males aged 18–34 years.

## Discussion

Using individual-level claims data, we found that the average per-person annual costs for the study OTC-like drugs among adults with representative chronic diseases were in the order of several hundred yen for acetaminophen and several thousand yen for NSAIDs, topical anti-inflammatory patches, second-generation antihistamines, and heparinoid preparations. These costs varied substantially by drug category, underlying chronic disease, age, and sex, suggesting that the financial burden may be disproportionately concentrated in specific strata. By applying stratum-specific estimates derived from the individual-level data to national population counts and aggregated claims data, we also estimated the national numbers of adults with representative chronic diseases who would need these drugs and the costs attributable to them. Across all study drugs and disease–age–sex strata, the estimated proportion (by number) of OTC-like drug users with representative chronic diseases was consistently <10%; in contrast, the maximum stratum-specific shares of total OTC-like drug costs attributable to these groups reached substantial levels (acetaminophen/NSAIDs/patches: around 50–60% for osteoarthritis, 10–20% for rheumatoid arthritis; antihistamines: around 80% for allergic rhinitis, ∼25% for urticaria, and ∼40% for atopic dermatitis; and heparinoids: ∼60% for atopic dermatitis/asteatosis). These patterns indicate that, although the number of patients with representative chronic diseases who would need OTC-like drugs may be small within the strata, their clinical needs—as reflected by cost concentration—may be substantial and compatible with long-term or recurrent use.

Several features of the stratum-specific patterns warrant consideration. First, the large variation in per-person annual costs by disease, age, and sex likely suggests differences in symptom burden, treatment duration, and prescribing practices, rather than a uniform “OTC-like drug user” profile. Second, the contrast between the small proportion of users (<10% in each stratum) and high-cost shares suggests that costs are concentrated among adults with high-intensity or long-term use, which is consistent with chronic, recurrent symptoms requiring repeated prescriptions.

A key issue in policy discussions on OTC-like drugs is how changes in coverage and cost-sharing affect the out-of-pocket burden for patients who rely on these medicines for chronic conditions. If coverage were to be removed entirely, the out-of-pocket impact would be compounded by two factors: (i) retail OTC drugs are typically several times more expensive than the corresponding OTC-like drugs, and in some cases the difference may be larger^7^; and (ii) OTC drugs require full out-of-pocket payment whereas patients generally pay only 10–30% coinsurance prescription because of the current reimbursement policy. Collectively, these factors could increase patient payments by several multiples of ten in some scenarios if the coverage of OTC-like drugs were be removed. According to the MHLW proposal released in December 2025, OTC-like drugs would remain covered, but a “special charge” equal to one quarter of the drug cost would be added, with coinsurance applied to the remaining three quarters^8^. Under this design, the effective patient share becomes 32.5% for 10% coinsurance, 40% for 20% coinsurance, and 47.5% for 30% coinsurance, corresponding to increases of 3.25-fold, 2.0-fold, and 1.6-fold, respectively, relative to the current coinsurance rates. To illustrate these effects, consider an example where the average annual total cost of antihistamines for allergic rhinitis in an adult aged ≥95 years is approximately 5000 yen (as shown in this study); currently, the patient currently pays 10% coinsurance (about 500 yen). If coverage were to be removed entirely and patients had to purchase OTC-like drugs—assuming that these are five times more expensive than their prescription-based counterparts—the annual out-of-pocket payment could rise to approximately 25000 yen. In contrast, under the proposed MHLW design, this payment would increase to about 1625 yen. The MHLW proposal also noted that special consideration would be given to certain subsets, including some patients with chronic diseases (e.g., malignant neoplasms and intractable diseases). Although it is uncertain whether the chronic diseases examined in this study would fall within the scope of any future special considerations, policymakers may wish to balance the expected per-person burden for patients requiring ongoing treatment against the potential national-level fiscal impact, drawing on the disease- and stratum-specific cost profiles presented here.

This study has several strengths. First, by using individual-level claims data, we described average per-person annual costs in detail across age–sex strata and across multiple chronic diseases linked to the indications for the study drugs. Second, the two-step approach—combining stratum-specific estimates from individual-level data with national population and aggregated claims costs—facilitated quantification of national figures and costs in a transparent and reproducible manner. In contrast, some limitations should be acknowledged. First, small cell values (generally <1000) in the NDB Open Data are masked, which can lead to modest underestimation of prescription costs; however, these masked components represent a small fraction of the total prescriptions and are therefore unlikely to materially change the overall magnitude of our national estimates. Second, the characteristics of individuals included in the DeSC database may differ from those of the overall Japanese population, which could affect the generalizability of stratum-specific estimates. Third, diagnoses were identified from claims codes and may not perfectly capture clinical indication or severity.

In conclusion, individual-level claims data showed that the per-person annual costs for OTC-like drugs varied widely by drug category and patient characteristics, and national estimation suggested that, within age–sex strata, small patient subgroups defined by representative chronic diseases may account for a disproportionately large share of the total costs of OTC-like drugs. These findings provide a quantitative baseline that may help inform ongoing policy discussions on cost-sharing for OTC-like drugs, including whether and how to operationalize special considerations for patients with chronic diseases.

## Author contributions

YK designed the research, collected the data, performed statistical analysis, and wrote the first draft. AS and AO designed the research and revised the first draft. HY revised the first draft and supervised the entire manuscript preparation. All authors have read and approved the final manuscript.

## Approval by institutional review board

This study protocol was approved by the Institutional Review Board of the Graduate School of Medicine of the University of Tokyo [approval number: 2021010NI (April 23, 2021)], and the study was conducted in accordance with the principles of the Helsinki Declaration.

## Data Availability

The NDB Open Data are available online at https://www.mhlw.go.jp/stf/seisakunitsuite/bunya/0000177182.html. The DeSC database is commercially available.

## Acknowledgements

The authors would like to thank Mr. Masayoshi Kurihara, a Health Information Manager at National Hospital Organization Tokyo National Hospital, for his helpful advice and information regarding OTC-like drugs. The authors used ChatGPT 5.2 (by OpenAI) and Gemini 3 (by Google) to enhance the readability of the English text during the preparation of this article. After using these tools, the authors reviewed and edited the content as needed and take full responsibility for the publication.

## Supplementary material

**Supplementary Table 1.** Acetaminophen use for osteoarthritis and rheumatoid arthritis by age-sex strata in the DeSC database.

| Disease and Age Category | Female | Male |
| --- | --- | --- |
| Osteoarthritis |  |  |
| 18–34 | 0.0% (72/214,705) | 0.0% (57/233,799) |
| 35–44 | 0.1% (207/143,278) | 0.1% (113/156,087) |
| 45–54 | 0.3% (587/186,727) | 0.2% (374/200,182) |
| 55–64 | 0.7% (1,629/220,303) | 0.4% (739/202,859) |
| 65–74 | 1.2% (7,231/618,707) | 0.6% (3,008/520,059) |
| 75–84 | 2.5% (22,419/904,800) | 1.2% (8,131/694,127) |
| 85–94 | 2.9% (13,241/462,011) | 1.7% (3,972/240,381) |
| ≥95 | 1.8% (1,248/68,486) | 1.3% (213/16,689) |
| Rheumatoid arthritis |  |  |
| 18–34 | 0.0% (52/214,705) | 0.0% (20/233,799) |
| 35–44 | 0.1% (92/143,278) | 0.0% (30/156,087) |
| 45–54 | 0.1% (220/186,727) | 0.0% (84/200,182) |
| 55–64 | 0.2% (400/220,303) | 0.1% (114/202,859) |
| 65–74 | 0.2% (1,407/618,707) | 0.1% (552/520,059) |
| 75–84 | 0.4% (3,565/904,800) | 0.2% (1,350/694,127) |
| 85–94 | 0.4% (1,738/462,011) | 0.2% (589/240,381) |
| ≥95 | 0.2% (122/68,486) | 0.1% (18/16,689) |

**Supplementary Table 2.** NSAID use for osteoarthritis and rheumatoid arthritis by age-sex strata in the DeSC database.

| Disease and Age Category | Female | Male |
| --- | --- | --- |
| Osteoarthritis |  |  |
| 18–34 | 0.0% (28/214,705) | 0.0% (26/233,799) |
| 35–44 | 0.1% (117/143,278) | 0.0% (69/156,087) |
| 45–54 | 0.2% (456/186,727) | 0.1% (293/200,182) |
| 55–64 | 0.6% (1,268/220,303) | 0.3% (606/202,859) |
| 65–74 | 1.0% (6,389/618,707) | 0.5% (2,492/520,059) |
| 75–84 | 2.0% (17,975/904,800) | 0.9% (6,293/694,127) |
| 85–94 | 1.8% (8,230/462,011) | 1.1% (2,537/240,381) |
| ≥95 | 0.7% (477/68,486) | 0.6% (96/16,689) |
| Rheumatoid arthritis |  |  |
| 18–34 | 0.0% (18/214,705) | 0.0% (13/233,799) |
| 35–44 | 0.0% (59/143,278) | 0.0% (19/156,087) |
| 45–54 | 0.1% (132/186,727) | 0.0% (48/200,182) |
| 55–64 | 0.1% (290/220,303) | 0.1% (102/202,859) |
| 65–74 | 0.2% (1,051/618,707) | 0.1% (404/520,059) |
| 75–84 | 0.3% (2,515/904,800) | 0.1% (903/694,127) |
| 85–94 | 0.2% (1,022/462,011) | 0.1% (292/240,381) |
| ≥95 | 0.1% (44/68,486) | 0.1% (10/16,689) |

NSAIDs: nonsteroidal anti-inflammatory drugs

**Supplementary Table 3.** Topical anti-inflammatory patches use for osteoarthritis and rheumatoid arthritis by age-sex strata in the DeSC database.

| Disease and Age Category | Female | Male |
| --- | --- | --- |
| **Osteoarthritis** |  |  |
| 18–34 | 0.0% (76/214,705) | 0.0% (73/233,799) |
| 35–44 | 0.2% (288/143,278) | 0.1% (229/156,087) |
| 45–54 | 0.7% (1,274/186,727) | 0.4% (832/200,182) |
| 55–64 | 1.6% (3,499/220,303) | 0.9% (1,763/202,859) |
| 65–74 | 3.4% (21,075/618,707) | 1.7% (8,752/520,059) |
| 75–84 | 7.0% (63,461/904,800) | 3.4% (23,577/694,127) |
| 85–94 | 6.8% (31,249/462,011) | 3.9% (9,479/240,381) |
| ≥95 | 3.1% (2,112/68,486) | 2.6% (439/16,689) |
| **Rheumatoid arthritis** |  |  |
| 18–34 | 0.0% (36/214,705) | 0.0% (8/233,799) |
| 35–44 | 0.1% (87/143,278) | 0.0% (27/156,087) |
| 45–54 | 0.1% (246/186,727) | 0.0% (96/200,182) |
| 55–64 | 0.3% (596/220,303) | 0.1% (182/202,859) |
| 65–74 | 0.4% (2,677/618,707) | 0.2% (1,001/520,059) |
| 75–84 | 0.8% (7,549/904,800) | 0.4% (2,620/694,127) |
| 85–94 | 0.7% (3,098/462,011) | 0.4% (852/240,381) |
| ≥95 | 0.2% (163/68,486) | 0.2% (30/16,689) |

**Supplementary Table 4.** Second-generation antihistamines use for allergic rhinitis, urticaria, and atopic dermatitis by age-sex strata in the DeSC database.

| Disease and Age Category | Female | Male |
| --- | --- | --- |
| Allergic rhinitis |  |  |
| 18–34 | 2.6% (5,594/214,705) | 1.9% (4,337/233,799) |
| 35–44 | 3.4% (4,892/143,278) | 2.2% (3,472/156,087) |
| 45–54 | 3.1% (5,864/186,727) | 2.0% (4,095/200,182) |
| 55–64 | 3.3% (7,176/220,303) | 2.3% (4,677/202,859) |
| 65–74 | 2.9% (17,663/618,707) | 2.3% (11,901/520,059) |
| 75–84 | 3.8% (34,486/904,800) | 3.5% (24,203/694,127) |
| 85–94 | 2.3% (10,536/462,011) | 2.7% (6,414/240,381) |
| ≥95 | 0.7% (495/68,486) | 1.2% (192/16,689) |
| Urticaria |  |  |
| 18–34 | 0.5% (1,137/214,705) | 0.3% (633/233,799) |
| 35–44 | 0.7% (970/143,278) | 0.3% (491/156,087) |
| 45–54 | 0.6% (1,204/186,727) | 0.3% (651/200,182) |
| 55–64 | 0.6% (1,407/220,303) | 0.4% (787/202,859) |
| 65–74 | 0.6% (3,585/618,707) | 0.4% (2,292/520,059) |
| 75–84 | 0.7% (6,559/904,800) | 0.6% (4,504/694,127) |
| 85–94 | 0.5% (2,121/462,011) | 0.5% (1,251/240,381) |
| ≥95 | 0.2% (110/68,486) | 0.3% (45/16,689) |
| Atopic dermatitis |  |  |
| 18–34 | 0.6% (1,238/214,705) | 0.5% (1,237/233,799) |
| 35–44 | 0.6% (912/143,278) | 0.5% (718/156,087) |
| 45–54 | 0.5% (1,016/186,727) | 0.3% (634/200,182) |
| 55–64 | 0.4% (806/220,303) | 0.2% (502/202,859) |
| 65–74 | 0.2% (1,530/618,707) | 0.2% (1,040/520,059) |
| 75–84 | 0.4% (3,768/904,800) | 0.4% (2,647/694,127) |
| 85–94 | 0.2% (1,076/462,011) | 0.3% (747/240,381) |
| ≥95 | 0.1% (54/68,486) | 0.2% (26/16,689) |

**Supplementary Table 5.** Heparinoid-containing topical preparations use for the combination of atopic dermatitis and asteatosis by age-sex strata in the DeSC database.

| Disease and Age Category | Female | Male |
| --- | --- | --- |
| Atopic dermatitis/Asteatosis |  |  |
| 18–34 | 0.4% (959/214,705) | 0.4% (1,007/233,799) |
| 35–44 | 0.4% (611/143,278) | 0.3% (523/156,087) |
| 45–54 | 0.3% (652/186,727) | 0.2% (447/200,182) |
| 55–64 | 0.2% (503/220,303) | 0.1% (297/202,859) |
| 65–74 | 0.2% (969/618,707) | 0.1% (711/520,059) |
| 75–84 | 0.3% (2,355/904,800) | 0.3% (1,846/694,127) |
| 85–94 | 0.2% (890/462,011) | 0.3% (680/240,381) |
| ≥95 | 0.1% (74/68,486) | 0.2% (35/16,689) |

**Supplementary Table 6.** Acetaminophen costs for osteoarthritis and rheumatoid arthritis by age-sex strata in the DeSC database.

| Disease and Age Category | Female | Male |
| --- | --- | --- |
| Osteoarthritis |  |  |
| 18–34 | 1.0% (15,150/1,559,505) | 0.8% (9,995/1,296,107) |
| 35–44 | 5.4% (66,257/1,223,122) | 2.4% (21,183/866,780) |
| 45–54 | 15.3% (264,024/1,730,590) | 10.7% (147,099/1,379,154) |
| 55–64 | 26.5% (698,554/2,632,457) | 17.9% (307,722/1,718,990) |
| 65–74 | 40.7% (3,452,431/8,482,698) | 25.5% (1,387,609/5,434,809) |
| 75–84 | 52.6% (12,230,917/23,271,633) | 30.5% (3,643,349/11,950,476) |
| 85–94 | 56.0% (8,816,672/15,730,395) | 38.9% (2,165,121/5,560,157) |
| ≥95 | 51.2% (691,332/1,350,130) | 30.1% (102,555/340,871) |
| Rheumatoid arthritis |  |  |
| 18–34 | 0.5% (8,218/1,559,505) | 0.2% (2,048/1,296,107) |
| 35–44 | 5.8% (70,439/1,223,122) | 1.6% (14,018/866,780) |
| 45–54 | 6.6% (114,908/1,730,590) | 3.6% (49,504/1,379,154) |
| 55–64 | 9.7% (255,952/2,632,457) | 2.4% (41,080/1,718,990) |
| 65–74 | 10.2% (866,241/8,482,698) | 5.3% (290,608/5,434,809) |
| 75–84 | 9.4% (2,193,276/23,271,633) | 6.5% (780,297/11,950,476) |
| 85–94 | 8.7% (1,364,583/15,730,395) | 6.1% (340,605/5,560,157) |
| ≥95 | 6.0% (80,743/1,350,130) | 1.8% (6,049/340,871) |

Costs are expressed in Japanese yen.

**Supplementary Table 7.** NSAID costs for osteoarthritis and rheumatoid arthritis by age-sex strata in the DeSC database.

| Disease and Age Category | Female | Male |
| --- | --- | --- |
| Osteoarthritis |  |  |
| 18–34 | 6.2% (41,192/668,484) | 4.1% (20,721/508,751) |
| 35–44 | 17.7% (197,960/1,118,860) | 6.5% (62,978/970,711) |
| 45–54 | 28.0% (751,651/2,687,452) | 17.9% (349,474/1,950,198) |
| 55–64 | 47.6% (2,853,833/5,997,571) | 30.2% (1,289,646/4,263,637) |
| 65–74 | 57.7% (14,857,447/25,737,350) | 38.5% (5,087,438/13,220,959) |
| 75–84 | 64.5% (51,793,316/80,331,893) | 46.9% (16,767,135/35,783,053) |
| 85–94 | 64.6% (27,866,131/43,144,318) | 50.9% (7,039,798/13,829,657) |
| ≥95 | 57.4% (1,698,895/2,962,056) | 44.7% (208,850/467,265) |
| Rheumatoid arthritis |  |  |
| 18–34 | 8.5% (56,629/668,484) | 2.8% (14,158/508,751) |
| 35–44 | 20.9% (233,337/1,118,860) | 5.2% (50,021/970,711) |
| 45–54 | 18.2% (489,791/2,687,452) | 5.8% (112,818/1,950,198) |
| 55–64 | 19.9% (1,194,187/5,997,571) | 6.6% (280,631/4,263,637) |
| 65–74 | 15.5% (3,991,235/25,737,350) | 11.5% (1,515,342/13,220,959) |
| 75–84 | 11.6% (9,304,692/80,331,893) | 9.2% (3,291,366/35,783,053) |
| 85–94 | 7.8% (3,362,447/43,144,318) | 5.9% (809,594/13,829,657) |
| ≥95 | 5.5% (161,476/2,962,056) | 6.3% (29,497/467,265) |

Costs are expressed in Japanese yen.
NSAIDs: nonsteroidal anti-inflammatory drugs

**Supplementary Table 8.** Costs of topical anti-inflammatory patches for osteoarthritis and rheumatoid arthritis by age-sex strata in the DeSC database.

| Disease and Age Category | Female | Male |
| --- | --- | --- |
| **Osteoarthritis** |  |  |
| 18–34 | 6.6% (132,759/2,011,304) | 4.9% (100,971/2,072,523) |
| 35–44 | 13.2% (561,727/4,245,575) | 10.8% (421,642/3,894,256) |
| 45–54 | 24.9% (2,651,069/10,643,510) | 19.4% (1,785,554/9,225,044) |
| 55–64 | 40.6% (9,916,081/24,404,825) | 29.9% (5,443,161/18,183,324) |
| 65–74 | 49.9% (74,075,180/148,537,761) | 33.0% (31,538,439/95,482,162) |
| 75–84 | 57.3% (332,376,463/579,930,491) | 39.0% (120,729,755/309,451,054) |
| 85–94 | 60.8% (164,140,456/269,924,399) | 45.3% (47,271,461/104,349,748) |
| ≥95 | 56.0% (8,870,214/15,840,061) | 46.4% (1,854,194/3,999,466) |
| **Rheumatoid arthritis** |  |  |
| 18–34 | 1.7% (33,348/2,011,304) | 0.2% (4,184/2,072,523) |
| 35–44 | 3.7% (155,448/4,245,575) | 2.0% (77,901/3,894,256) |
| 45–54 | 5.8% (612,807/10,643,510) | 1.9% (179,461/9,225,044) |
| 55–64 | 8.2% (1,989,624/24,404,825) | 2.0% (369,923/18,183,324) |
| 65–74 | 6.5% (9,697,336/148,537,761) | 3.7% (3,490,139/95,482,162) |
| 75–84 | 6.4% (37,203,379/579,930,491) | 3.9% (12,155,224/309,451,054) |
| 85–94 | 5.6% (15,178,625/269,924,399) | 3.7% (3,820,480/104,349,748) |
| ≥95 | 4.0% (639,383/15,840,061) | 2.6% (102,768/3,999,466) |

Costs are expressed in Japanese yen.

**Supplementary Table 9.** Costs of second-generation antihistamines for allergic rhinitis, urticaria, and atopic dermatitis by age-sex strata in the DeSC database.

| Disease and Age Category | Female | Male |
| --- | --- | --- |
| **Allergic rhinitis** |  |  |
| 18–34 | 75.9% (9,240,003/12,176,037) | 66.3% (7,466,404/11,262,762) |
| 35–44 | 78.5% (11,041,876/14,070,832) | 71.8% (7,784,601/10,844,474) |
| 45–54 | 77.4% (16,899,046/21,820,533) | 70.8% (11,882,611/16,782,870) |
| 55–64 | 77.8% (21,069,622/27,081,094) | 72.6% (14,629,945/20,142,456) |
| 65–74 | 75.2% (51,383,188/68,347,935) | 69.2% (36,237,666/52,348,692) |
| 75–84 | 78.0% (129,639,857/166,304,537) | 72.4% (91,180,512/125,866,164) |
| 85–94 | 71.1% (47,526,514/66,882,039) | 69.5% (28,073,868/40,407,096) |
| ≥95 | 56.6% (2,409,668/4,257,716) | 58.9% (943,061/1,600,991) |
| **Urticaria** |  |  |
| 18–34 | 20.1% (2,442,503/12,176,037) | 13.3% (1,502,906/11,262,762) |
| 35–44 | 23.3% (3,283,478/14,070,832) | 15.6% (1,692,605/10,844,474) |
| 45–54 | 23.2% (5,061,632/21,820,533) | 17.5% (2,930,105/16,782,870) |
| 55–64 | 19.0% (5,141,348/27,081,094) | 15.6% (3,148,256/20,142,456) |
| 65–74 | 18.6% (12,736,478/68,347,935) | 17.3% (9,049,371/52,348,692) |
| 75–84 | 14.9% (24,734,296/166,304,537) | 15.3% (19,242,524/125,866,164) |
| 85–94 | 13.2% (8,829,788/66,882,039) | 13.3% (5,392,273/40,407,096) |
| ≥95 | 11.2% (475,352/4,257,716) | 9.7% (154,899/1,600,991) |
| **Atopic dermatitis** |  |  |
| 18–34 | 27.6% (3,356,777/12,176,037) | 37.3% (4,204,428/11,262,762) |
| 35–44 | 24.5% (3,445,960/14,070,832) | 26.0% (2,822,744/10,844,474) |
| 45–54 | 20.1% (4,383,215/21,820,533) | 21.9% (3,679,341/16,782,870) |
| 55–64 | 13.6% (3,676,090/27,081,094) | 15.0% (3,024,803/20,142,456) |
| 65–74 | 9.1% (6,190,649/68,347,935) | 9.8% (5,125,252/52,348,692) |
| 75–84 | 10.2% (16,901,944/166,304,537) | 11.7% (14,774,097/125,866,164) |
| 85–94 | 8.3% (5,535,998/66,882,039) | 10.2% (4,131,427/40,407,096) |
| ≥95 | 6.7% (286,416/4,257,716) | 8.2% (132,001/1,600,991) |

Costs are expressed in Japanese yen.

**Supplementary Table 10.** Costs of heparinoid-containing topical preparations for the combination of atopic dermatitis and asteatosis by age-sex strata in the DeSC database.

| Disease and Age Category | Female | Male |
| --- | --- | --- |
| Atopic dermatitis/Asteatosis |  |  |
| 18–34 | 42.9% (2,386,483/5,557,189) | 58.2% (3,270,876/5,623,019) |
| 35–44 | 37.6% (1,979,049/5,267,266) | 53.4% (1,889,550/3,540,361) |
| 45–54 | 30.4% (2,532,713/8,328,648) | 38.9% (1,849,761/4,758,593) |
| 55–64 | 19.4% (1,715,757/8,827,771) | 23.0% (1,172,887/5,102,686) |
| 65–74 | 10.4% (2,475,004/23,735,331) | 13.7% (2,487,583/18,107,982) |
| 75–84 | 10.4% (8,548,966/82,568,730) | 14.2% (8,348,290/58,901,226) |
| 85–94 | 7.1% (3,086,039/43,687,261) | 11.1% (2,461,510/22,204,105) |
| ≥95 | 8.9% (503,791/5,644,204) | 6.4% (95,963/1,491,701) |

Costs are expressed in Japanese yen.

**Supplementary Table 11.** Estimated national proportion of adults needing acetaminophen for osteoarthritis and rheumatoid arthritis by age-sex strata.

| Disease and Age Category | Female | Male |
| --- | --- | --- |
| **Osteoarthritis** |  |  |
| 18–34 | 0.0% (3,477/10,367,000) | 0.0% (2,674/10,970,000) |
| 35–44 | 0.1% (10,519/7,281,000) | 0.1% (5,452/7,531,000) |
| 45–54 | 0.3% (29,167/9,278,000) | 0.2% (17,728/9,489,000) |
| 55–64 | 0.7% (58,593/7,924,000) | 0.4% (28,644/7,863,000) |
| 65–74 | 1.2% (98,407/8,420,000) | 0.6% (44,704/7,729,000) |
| 75–84 | 2.5% (186,974/7,546,000) | 1.2% (68,222/5,824,000) |
| 85–94 | 2.9% (114,495/3,995,000) | 1.7% (33,560/2,031,000) |
| ≥95 | 1.8% (10,114/555,000) | 1.3% (1,685/132,000) |
| **Rheumatoid arthritis** |  |  |
| 18–34 | 0.0% (2,511/10,367,000) | 0.0% (938/10,970,000) |
| 35–44 | 0.1% (4,675/7,281,000) | 0.0% (1,447/7,531,000) |
| 45–54 | 0.1% (10,931/9,278,000) | 0.0% (3,982/9,489,000) |
| 55–64 | 0.2% (14,387/7,924,000) | 0.1% (4,419/7,863,000) |
| 65–74 | 0.2% (19,148/8,420,000) | 0.1% (8,204/7,729,000) |
| 75–84 | 0.4% (29,732/7,546,000) | 0.2% (11,327/5,824,000) |
| 85–94 | 0.4% (15,028/3,995,000) | 0.2% (4,977/2,031,000) |
| ≥95 | 0.2% (989/555,000) | 0.1% (142/132,000) |

**Supplementary Table 12.** Estimated national proportion of adults needing NSAIDs for osteoarthritis and rheumatoid arthritis by age-sex strata.

| Disease and Age Category | Female | Male |
| --- | --- | --- |
| **Osteoarthritis** |  |  |
| 18–34 | 0.0% (1,352/10,367,000) | 0.0% (1,220/10,970,000) |
| 35–44 | 0.1% (5,946/7,281,000) | 0.0% (3,329/7,531,000) |
| 45–54 | 0.2% (22,658/9,278,000) | 0.1% (13,889/9,489,000) |
| 55–64 | 0.6% (45,608/7,924,000) | 0.3% (23,489/7,863,000) |
| 65–74 | 1.0% (86,948/8,420,000) | 0.5% (37,036/7,729,000) |
| 75–84 | 2.0% (149,911/7,546,000) | 0.9% (52,801/5,824,000) |
| 85–94 | 1.8% (71,165/3,995,000) | 1.1% (21,435/2,031,000) |
| ≥95 | 0.7% (3,866/555,000) | 0.6% (759/132,000) |
| **Rheumatoid arthritis** |  |  |
| 18–34 | 0.0% (869/10,367,000) | 0.0% (610/10,970,000) |
| 35–44 | 0.0% (2,998/7,281,000) | 0.0% (917/7,531,000) |
| 45–54 | 0.1% (6,559/9,278,000) | 0.0% (2,275/9,489,000) |
| 55–64 | 0.1% (10,431/7,924,000) | 0.1% (3,954/7,863,000) |
| 65–74 | 0.2% (14,303/8,420,000) | 0.1% (6,004/7,729,000) |
| 75–84 | 0.3% (20,975/7,546,000) | 0.1% (7,577/5,824,000) |
| 85–94 | 0.2% (8,837/3,995,000) | 0.1% (2,467/2,031,000) |
| ≥95 | 0.1% (357/555,000) | 0.1% (79/132,000) |

NSAIDs: nonsteroidal anti-inflammatory drugs

**Supplementary Table 13.** Estimated national proportion of adults needing topical anti-inflammatory patches for osteoarthritis and rheumatoid arthritis by age-sex strata.

| Disease and Age Category | Female | Male |
| --- | --- | --- |
| Osteoarthritis |  |  |
| 18–34 | 0.0% (3,670/10,367,000) | 0.0% (3,425/10,970,000) |
| 35–44 | 0.2% (14,635/7,281,000) | 0.1% (11,049/7,531,000) |
| 45–54 | 0.7% (63,302/9,278,000) | 0.4% (39,438/9,489,000) |
| 55–64 | 1.6% (125,854/7,924,000) | 0.9% (68,335/7,863,000) |
| 65–74 | 3.4% (286,810/8,420,000) | 1.7% (130,070/7,729,000) |
| 75–84 | 7.0% (529,262/7,546,000) | 3.4% (197,820/5,824,000) |
| 85–94 | 6.8% (270,209/3,995,000) | 3.9% (80,089/2,031,000) |
| ≥95 | 3.1% (17,115/555,000) | 2.6% (3,472/132,000) |
| Rheumatoid arthritis |  |  |
| 18–34 | 0.0% (1,738/10,367,000) | 0.0% (375/10,970,000) |
| 35–44 | 0.1% (4,421/7,281,000) | 0.0% (1,303/7,531,000) |
| 45–54 | 0.1% (12,223/9,278,000) | 0.0% (4,551/9,489,000) |
| 55–64 | 0.3% (21,437/7,924,000) | 0.1% (7,054/7,863,000) |
| 65–74 | 0.4% (36,431/8,420,000) | 0.2% (14,877/7,729,000) |
| 75–84 | 0.8% (62,958/7,546,000) | 0.4% (21,983/5,824,000) |
| 85–94 | 0.7% (26,788/3,995,000) | 0.4% (7,199/2,031,000) |
| ≥95 | 0.2% (1,321/555,000) | 0.2% (237/132,000) |

**Supplementary Table 14.** Estimated national proportion of adults needing second-generation antihistamines for allergic rhinitis, urticaria, and atopic dermatitis by age-sex strata.

| Disease and Age Category | Female | Male |
| --- | --- | --- |
| Allergic rhinitis |  |  |
| 18–34 | 2.6% (270,105/10,367,000) | 1.9% (203,495/10,970,000) |
| 35–44 | 3.4% (248,598/7,281,000) | 2.2% (167,520/7,531,000) |
| 45–54 | 3.1% (291,368/9,278,000) | 2.0% (194,111/9,489,000) |
| 55–64 | 3.3% (258,111/7,924,000) | 2.3% (181,285/7,863,000) |
| 65–74 | 2.9% (240,376/8,420,000) | 2.3% (176,870/7,729,000) |
| 75–84 | 3.8% (287,612/7,546,000) | 3.5% (203,073/5,824,000) |
| 85–94 | 2.3% (91,105/3,995,000) | 2.7% (54,192/2,031,000) |
| ≥95 | 0.7% (4,011/555,000) | 1.2% (1,519/132,000) |
| Urticaria |  |  |
| 18–34 | 0.5% (54,900/10,367,000) | 0.3% (29,701/10,970,000) |
| 35–44 | 0.7% (49,293/7,281,000) | 0.3% (23,690/7,531,000) |
| 45–54 | 0.6% (59,824/9,278,000) | 0.3% (30,859/9,489,000) |
| 55–64 | 0.6% (50,608/7,924,000) | 0.4% (30,505/7,863,000) |
| 65–74 | 0.6% (48,788/8,420,000) | 0.4% (34,063/7,729,000) |
| 75–84 | 0.7% (54,702/7,546,000) | 0.6% (37,790/5,824,000) |
| 85–94 | 0.5% (18,340/3,995,000) | 0.5% (10,570/2,031,000) |
| ≥95 | 0.2% (891/555,000) | 0.3% (356/132,000) |
| Atopic dermatitis |  |  |
| 18–34 | 0.6% (59,777/10,367,000) | 0.5% (58,041/10,970,000) |
| 35–44 | 0.6% (46,345/7,281,000) | 0.5% (34,643/7,531,000) |
| 45–54 | 0.5% (50,483/9,278,000) | 0.3% (30,053/9,489,000) |
| 55–64 | 0.4% (28,991/7,924,000) | 0.2% (19,458/7,863,000) |
| 65–74 | 0.2% (20,822/8,420,000) | 0.2% (15,456/7,729,000) |
| 75–84 | 0.4% (31,425/7,546,000) | 0.4% (22,209/5,824,000) |
| 85–94 | 0.2% (9,304/3,995,000) | 0.3% (6,311/2,031,000) |
| ≥95 | 0.1% (438/555,000) | 0.2% (206/132,000) |

**Supplementary Table 15.**
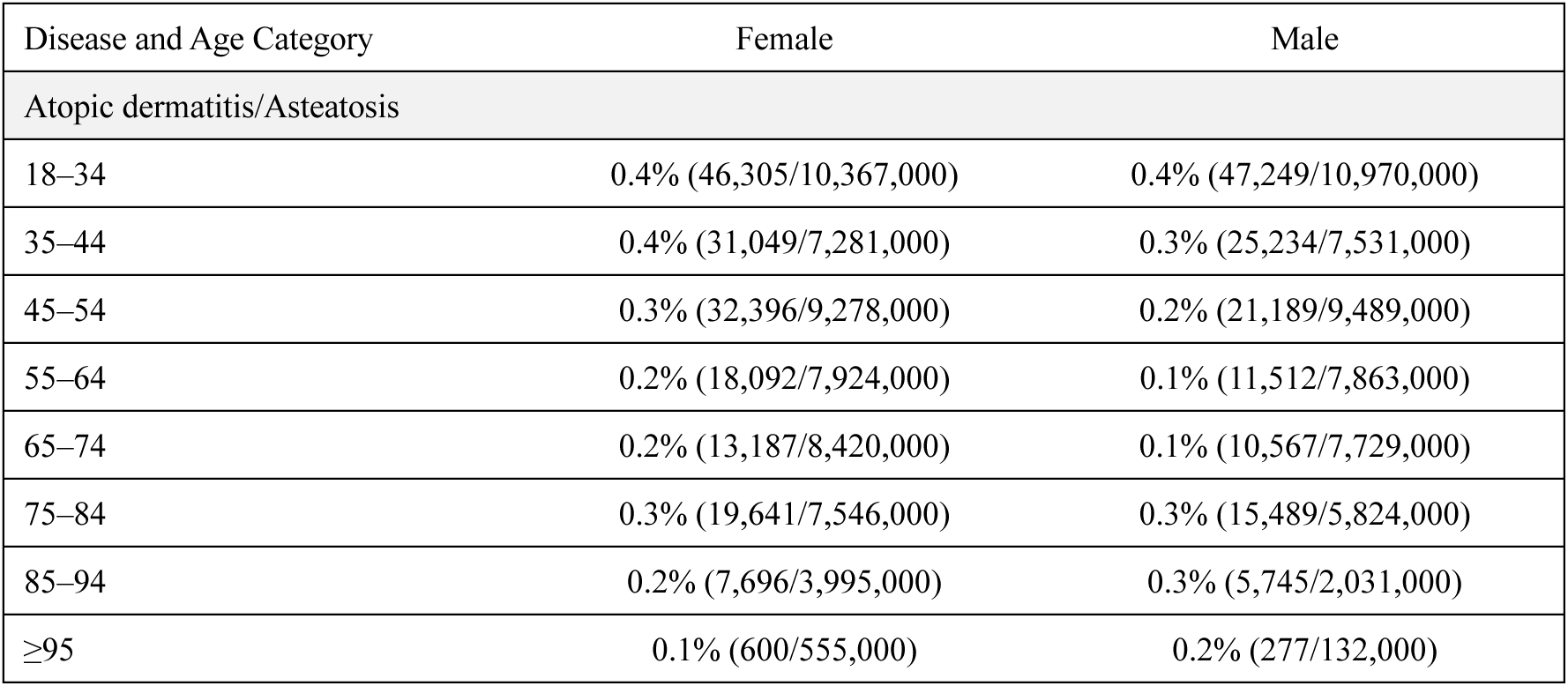
Estimated national proportion of adults needing heparinoid-containing topical preparations for the combination of atopic dermatitis and asteatosis by age-sex strata.

**Supplementary Table 16.** Estimated national costs of acetaminophen for osteoarthritis and rheumatoid arthritis by age-sex strata.

| Disease and Age Category | Female | Male |
| --- | --- | --- |
| **Osteoarthritis** |  |  |
| 18–34 | 1.0% (8,151,303/839,075,746) | 0.8% (5,074,822/658,080,244) |
| 35–44 | 5.4% (28,283,779/522,126,149) | 2.4% (8,077,551/330,522,562) |
| 45–54 | 15.3% (99,893,843/654,771,105) | 10.7% (45,505,717/426,647,302) |
| 55–64 | 26.5% (187,639,247/707,106,751) | 17.9% (88,251,468/492,988,450) |
| 65–74 | 40.7% (414,036,397/1,017,296,426) | 25.5% (178,300,069/698,342,847) |
| 75–84 | 52.6% (962,251,137/1,830,864,792) | 30.5% (275,620,882/904,058,527) |
| 85–94 | 56.0% (980,192,921/1,748,825,614) | 38.9% (212,061,280/544,585,734) |
| ≥95 | 51.2% (139,717,075/272,859,081) | 30.1% (14,192,510/47,172,882) |
| **Rheumatoid arthritis** |  |  |
| 18–34 | 0.5% (4,421,611/839,075,746) | 0.2% (1,039,843/658,080,244) |
| 35–44 | 5.8% (30,068,991/522,126,149) | 1.6% (5,345,376/330,522,562) |
| 45–54 | 6.6% (43,475,600/654,771,105) | 3.6% (15,314,278/426,647,302) |
| 55–64 | 9.7% (68,751,507/707,106,751) | 2.4% (11,781,317/492,988,450) |
| 65–74 | 10.2% (103,884,858/1,017,296,426) | 5.3% (37,341,518/698,342,847) |
| 75–84 | 9.4% (172,553,074/1,830,864,792) | 6.5% (59,029,796/904,058,527) |
| 85–94 | 8.7% (151,707,424/1,748,825,614) | 6.1% (33,360,321/544,585,734) |
| ≥95 | 6.0% (16,318,029/272,859,081) | 1.8% (837,117/47,172,882) |

Costs are expressed in Japanese yen.

**Supplementary Table 17.** Estimated national costs of NSAIDs for osteoarthritis and rheumatoid arthritis by age-sex strata.

| Disease and Age Category | Female | Male |
| --- | --- | --- |
| Osteoarthritis |  |  |
| 18–34 | 6.2% (13,954,521/226,460,821) | 4.1% (7,675,794/188,459,423) |
| 35–44 | 17.7% (52,219,569/295,142,389) | 6.5% (15,830,241/243,999,320) |
| 45–54 | 28.0% (218,299,491/780,507,713) | 17.9% (99,419,127/554,796,589) |
| 55–64 | 47.6% (553,078,203/1,162,340,540) | 30.2% (246,473,453/814,854,104) |
| 65–74 | 57.7% (1,112,193,960/1,926,638,219) | 38.5% (439,356,950/1,141,777,102) |
| 75–84 | 64.5% (1,850,609,112/2,870,311,165) | 46.9% (580,313,777/1,238,458,367) |
| 85–94 | 64.6% (1,165,106,731/1,803,900,775) | 50.9% (270,993,478/532,365,680) |
| ≥95 | 57.4% (108,320,054/188,858,090) | 44.7% (14,279,724/31,948,361) |
| Rheumatoid arthritis |  |  |
| 18–34 | 8.5% (19,184,079/226,460,821) | 2.8% (5,244,626/188,459,423) |
| 35–44 | 20.9% (61,551,615/295,142,389) | 5.2% (12,573,351/243,999,320) |
| 45–54 | 18.2% (142,248,365/780,507,713) | 5.8% (32,094,711/554,796,589) |
| 55–64 | 19.9% (231,435,687/1,162,340,540) | 6.6% (53,633,394/814,854,104) |
| 65–74 | 15.5% (298,774,578/1,926,638,219) | 11.5% (130,866,664/1,141,777,102) |
| 75–84 | 11.6% (332,462,741/2,870,311,165) | 9.2% (113,914,812/1,238,458,367) |
| 85–94 | 7.8% (140,586,780/1,803,900,775) | 5.9% (31,164,913/532,365,680) |
| ≥95 | 5.5% (10,295,568/188,858,090) | 6.3% (2,016,802/31,948,361) |

Costs are expressed in Japanese yen.
NSAIDs: nonsteroidal anti-inflammatory drugs

**Supplementary Table 18.** Estimated national costs of topical anti-inflammatory patches for osteoarthritis and rheumatoid arthritis by age-sex strata.

| Disease and Age Category | Female | Male |
| --- | --- | --- |
| **Osteoarthritis** |  |  |
| 18–34 | 6.6% (53,692,904/813,449,577) | 4.9% (38,257,151/785,263,355) |
| 35–44 | 13.2% (166,218,748/1,256,293,821) | 10.8% (108,112,915/998,523,319) |
| 45–54 | 24.9% (790,811,089/3,174,947,818) | 19.4% (456,291,244/2,357,423,410) |
| 55–64 | 40.6% (2,088,219,929/5,139,393,468) | 29.9% (1,178,654,986/3,937,393,271) |
| 65–74 | 49.9% (5,729,652,395/11,489,269,929) | 33.0% (2,566,445,088/7,769,874,904) |
| 75–84 | 57.3% (11,440,154,428/19,960,782,767) | 39.0% (4,107,356,248/10,527,858,026) |
| 85–94 | 60.8% (6,907,293,510/11,358,851,406) | 45.3% (1,878,504,852/4,146,719,897) |
| ≥95 | 56.0% (559,060,098/998,346,382) | 46.4% (96,442,592/208,025,086) |
| **Rheumatoid arthritis** |  |  |
| 18–34 | 1.7% (13,487,228/813,449,577) | 0.2% (1,585,286/785,263,355) |
| 35–44 | 3.7% (45,998,095/1,256,293,821) | 2.0% (19,974,538/998,523,319) |
| 45–54 | 5.8% (182,799,682/3,174,947,818) | 1.9% (45,860,547/2,357,423,410) |
| 55–64 | 8.2% (418,993,399/5,139,393,468) | 2.0% (80,102,644/3,937,393,271) |
| 65–74 | 6.5% (750,080,721/11,489,269,929) | 3.7% (284,010,572/7,769,874,904) |
| 75–84 | 6.4% (1,280,513,058/19,960,782,767) | 3.9% (413,533,807/10,527,858,026) |
| 85–94 | 5.6% (638,740,872/11,358,851,406) | 3.7% (151,820,783/4,146,719,897) |
| ≥95 | 4.0% (40,298,185/998,346,382) | 2.6% (5,345,294/208,025,086) |

Costs are expressed in Japanese yen.

**Supplementary Table 19.** Estimated national costs of second-generation antihistamines for allergic rhinitis, urticaria, and atopic dermatitis by age-sex strata.

| Disease and Age Category | Female | Male |
| --- | --- | --- |
| **Allergic rhinitis** |  |  |
| 18–34 | 75.9% (7,210,462,617/9,501,605,098) | 66.3% (5,105,394,057/7,701,276,034) |
| 35–44 | 78.5% (6,166,068,134/7,857,515,228) | 71.8% (3,426,822,939/4,773,795,377) |
| 45–54 | 77.4% (8,284,068,470/10,696,626,864) | 70.8% (4,415,362,330/6,236,209,533) |
| 55–64 | 77.8% (7,314,187,137/9,401,031,940) | 72.6% (4,559,264,677/6,277,179,316) |
| 65–74 | 75.2% (6,305,050,347/8,386,734,807) | 69.2% (4,458,940,691/6,441,356,153) |
| 75–84 | 78.0% (5,787,900,565/7,424,831,730) | 72.4% (4,207,170,476/5,807,605,129) |
| 85–94 | 71.1% (2,504,255,280/3,524,131,800) | 69.5% (1,537,134,852/2,212,418,878) |
| ≥95 | 56.6% (245,035,802/432,961,244) | 58.9% (85,981,404/145,966,649) |
| **Urticaria** |  |  |
| 18–34 | 20.1% (1,906,014,162/9,501,605,098) | 13.3% (1,027,660,352/7,701,276,034) |
| 35–44 | 23.3% (1,833,578,738/7,857,515,228) | 15.6% (745,093,761/4,773,795,377) |
| 45–54 | 23.2% (2,481,258,768/10,696,626,864) | 17.5% (1,088,773,776/6,236,209,533) |
| 55–64 | 19.0% (1,784,786,714/9,401,031,940) | 15.6% (981,120,050/6,277,179,316) |
| 65–74 | 18.6% (1,562,848,437/8,386,734,807) | 17.3% (1,113,499,103/6,441,356,153) |
| 75–84 | 14.9% (1,104,287,286/7,424,831,730) | 15.3% (887,871,510/5,807,605,129) |
| 85–94 | 13.2% (465,256,998/3,524,131,800) | 13.3% (295,244,345/2,212,418,878) |
| ≥95 | 11.2% (48,337,887/432,961,244) | 9.7% (14,122,558/145,966,649) |
| **Atopic dermatitis** |  |  |
| 18–34 | 27.6% (2,619,470,478/9,501,605,098) | 37.3% (2,874,912,974/7,701,276,034) |
| 35–44 | 24.5% (1,924,312,875/7,857,515,228) | 26.0% (1,242,586,986/4,773,795,377) |
| 45–54 | 20.1% (2,148,692,487/10,696,626,864) | 21.9% (1,367,176,259/6,236,209,533) |
| 55–64 | 13.6% (1,276,131,588/9,401,031,940) | 15.0% (942,647,253/6,277,179,316) |
| 65–74 | 9.1% (759,632,774/8,386,734,807) | 9.8% (630,647,534/6,441,356,153) |
| 75–84 | 10.2% (754,604,128/7,424,831,730) | 11.7% (681,693,306/5,807,605,129) |
| 85–94 | 8.3% (291,701,433/3,524,131,800) | 10.2% (226,208,958/2,212,418,878) |
| ≥95 | 6.7% (29,125,246/432,961,244) | 8.2% (12,034,886/145,966,649) |

Costs are expressed in Japanese yen.

**Supplementary Table 20.** Estimated national costs of heparinoid-containing topical preparations for the combination of atopic dermatitis and asteatosis by age-sex strata.

| Disease and Age Category | Female | Male |
| --- | --- | --- |
| Atopic dermatitis/Asteatosis |  |  |
| 18–34 | 42.9% (2,200,519,393/5,124,152,222) | 58.2% (2,623,794,036/4,510,609,303) |
| 35–44 | 37.6% (1,031,245,050/2,744,672,815) | 53.4% (866,091,975/1,622,755,816) |
| 45–54 | 30.4% (1,121,148,254/3,686,816,929) | 38.9% (618,780,870/1,591,841,496) |
| 55–64 | 19.4% (596,072,795/3,066,864,439) | 23.0% (343,055,145/1,492,473,431) |
| 65–74 | 10.4% (318,357,853/3,053,057,291) | 13.7% (295,806,947/2,153,281,665) |
| 75–84 | 10.4% (392,895,401/3,794,713,218) | 14.2% (400,516,977/2,825,841,097) |
| 85–94 | 7.1% (179,818,160/2,545,581,216) | 11.1% (156,213,769/1,409,129,729) |
| ≥95 | 8.9% (43,219,431/484,207,309) | 6.4% (8,914,791/138,576,352) |

Costs are expressed in Japanese yen.

